# What happened to health labour markets during COVID-19? Insights from a survey of medical doctors in Brazil

**DOI:** 10.1101/2023.05.03.23289458

**Authors:** Bruno Luciano Carneiro Alves de Oliveira, Mário Scheffer, Alex Cassenote, Giuliano Russo

## Abstract

**Background:** Limited evidence exists on impacts and adaptations of global health markets during COVID-19. We examined physicians’ perceptions of changing employment opportunities in Brazil, to gain an insight into labour markets in low- and middle-income countries (LMICs) during the pandemic.

**Methods:** We conducted secondary analysis of a dataset from a representative cross-sectional survey of 1,183 physicians in São Paulo and Maranhão states in Brazil. We estimated prevalence and 95% Confidence Intervals (CI) for proxy variables of demand and supply of doctors, and prices of medical services for facilities of practice in the two States, stratified by public, private, and dual practice physicians.

**Results:** Most doctors reported increased job opportunities in the public sector (59.0%, 95% CI 56.1-61.9), particularly in Maranhão state (66.4%, 95% CI 62.3-70.3). For the private sector, increased opportunities were reported only in large private hospitals (51.4%, 95% CI 48.4-54.4), but not in smaller clinics. We recorded perceptions of slight increases in availability of doctors in Maranhão, particularly in the public sector (54.1%, 95 CI 45.7-62.3). Younger doctors recounted increased vacancies in the public sector (64%, 95 CI 58.1-68.1); older doctors only in walk-in clinics in Maranhão (47.5%, 95 CI 39.9-55.1). Those working directly with COVID-19 saw opportunities in public hospitals (65%, 95 CI 62.3-68.4), and in large private ones (55%, 95 CI 51.8-59.1)

**Conclusions:** Our findings suggest that health labour markets in (LMICs) may not necessarily shrink during epidemics, and that impacts will depend on the balance of public and private services in national health systems.

**Key messages:** 

**What is already known on this topi:** Health labour markets are believed to shrink during epidemics, with fewer services and jobs available because of lockdowns and reduced demand.

**What this study adds:** The doctors we surveyed in Brazil noticed increased job opportunities in the public sector during COVID-19, particularly in Maranhão state. For the private sector, increased vacancies were reported in large private hospitals but not in smaller clinics.

**How this study might affect research, practice or policy:** The complementary roles of health markets and publicly or privately funded systems during a health emergency might need re-examining to improve pandemic preparedness in LMICs.

## Introduction and background

The COVID-19 pandemic and the ensuing economic crisis has had an unparalleled impact on population health ^1^, national economies ^2^, and working modalities ^3^. A recent OECD report presents evidence on the ways labour markets have changed due to pandemic-related lockdown measures, as businesses were disrupted and workers furloughed or laid off ^4^.

The economic literature has so far suggested that health labour markets are ‘recession proof’, that is, one of the few sectors to hold up during economic slowdowns ^5^. Scholars have argued this may be due to an inelastic demand for healthcare services, and to increasing health needs during recessions ^6^. Evidence from the US in fact showed increasing employment opportunities in the healthcare industry during the 2008 financial crisis, particularly for nurses ^7^. Studies from Canada and Australia concluded that labour markets for physicians were largely unaffected during the ensuing recession ^8,9^. However, in those countries with publicly funded health systems, government-driven austerity measures did reduce health sector resources and shrink health labour markets, particularly in Europe ^10–12^.

There is a growing body of literature highlighting the role of labour markets in the provision of healthcare services in both high-income countries and LMICs ^13,14^, which tends to advocate for the use of Health Labour Market Analysis to design effective health policies in such contexts ^15,16^.

During COVID-19, research has shown that global labour health markets experienced a multi-layered crisis amplified by the combination of lockdown measures and reduced worker mobility ^17^. In the first year of the pandemic, studies from the US argued that health sector unemployment rose because of pay cuts and redundancies, as patients were delaying seeking treatment, and hospitals were focussing on COVID-19 care, which “is not where the money is” ^18^. Other US scholars ^19^ noticed that unemployment in the healthcare industry increased less than in other sectors during COVID-19; while less specialised jobs (such as non-healthcare hospital workers and therapists) were badly affected, employment opportunities for physicians barely shifted. As many patients were moved to government-funded schemes, new “price sensitivities” were found in the demand for healthcare service, as customers would no longer be insulated from costs ^20^.

The evidence from outside the US is less consolidated. Analysis of labour statistics and employment censuses from the UK market ^21^ show that healthcare employment declined suddenly in 2020 only to bounce back the year after, with dentists and nurses among the worst affected professions. Another British study looking at job ads during the pandemic ^22^ found more care work and nursing vacancies than in any other sector during lockdowns. Similar findings were reported for Serbia, where COVID-19 seems to have entrenched a continuous mismatch between supply and demand for physicians and nurses ^23^. An online survey study from Iran ^24^ suggested that healthcare employment would have become less attractive because of COVID-19, as more health workers consider leaving. At the height of the first wave of the pandemic, there were reports ^25^ that Mexico, South Africa, and Zambia were recruiting doctors from abroad, as a surge in the demand for COVID-19 care was expected.

Brazil has been one of the world’s most affected countries, with over 700,000 COVID-19 related deaths; its economy contracted by 3.9% in 2020, although it rebounded by 4.6% and 2.6% in the following years ^26^. The country’s unemployment rate is currently at 9%, and the International Monetary Fund estimates that approximately 12 million jobs have been lost as a consequence of the pandemic, with a disproportionate impact among the lower income groups ^27^. Past economic recessions historically had little repercussions for physician employment, as in Brazil demand has always outstripped supply ^28^. But a recent study showed that public sector physicians’ workload and earnings increased in Brazil during the first two years of the pandemic, while those of private doctors suffered ^29^.

Brazil’s healthcare system comprises a publicly funded Unified Health System (*Sistema Único de Saúde*, SUS) and a multiplicity of privately financed sub-sectors, including large, comprehensive private hospitals and smaller, outpatient care surgeries ^30^. In the last 20 years, low-cost walk-in clinics in urban areas (*Clínicas Populares*, or People’s Clinics) have started to provide out-of-pocket private services, mostly outpatient in nature ^31,32^. Within SUS, provision of free-at-the-point-of-care services is often outsourced to private entities (social health organisations) that manage public hospitals and contract their own staff, including doctors. Private funds account for more than half (54%) of Brazil’s health spending, including out-of-pocket medicines and private insurance premiums ^33^. Access to such private services is funded through employment-related health insurance plans, with 24.2% of Brazil’s population owning such private plans ^34^.

This paper uses physicians’ perceptions of changing employment opportunities in Brazil to gain an insight into health labour markets during COVID-19 in LMICs. The aim of this study is to contribute to the existing body of work on health labour markets in mixed health systems across the world; this would provide an evidence base for policies to mitigate the effects of future shocks on health workforces.

### Methods

#### Methodological approach

As part of a wider study on the health workforce in two Brazilian states ^35^, we conducted secondary analysis of a dataset from a representative cross-sectional survey on physicians’ perceptions of the impact of COVID-19 on their health, earnings, and work routines ^29,36,37^. Workers’ perceptions have been used before in the economic literature to explore labour market dynamics in high-income contexts38.

In our survey, doctors were asked whether they had observed in the past two years:

(a) an increase or a decline of job opportunities and vacancies in their workplace; (b) any change in the availability of doctors; and (c) any change in the remuneration of a 12-hour shift in their institution’s Accidents and Emergency (A&E) ward (see the Survey Questionnaire in Supplementary Information 1). We interpreted the reported changes in (a) as proxies for changes in demand for doctors; responses on (b) as proxies for supply; and responses on (c) as a proxy for changes in the level of prices for healthcare services.

As our doctors worked in either public or private facilities, or both, we considered the doctors’ perceptions of changes in their own sector of employment as particularly accurate. As a way of validating responses, we triangulated the perceptions of the entire sample with those from doctors working specifically in such sectors. We outline the limitations of such an approach in the Discussion.

#### Study settings and data collection

The original survey was carried out in one rich state (São Paulo) and one less developed one (Maranhão) in Brazil, with a view to capture the differential effects of economic recessions in diverse health markets ^34^. São Paulo state is home to more than 46.6M people, has one of the country’s highest per capita incomes, and 38% of its population is covered by private health schemes. The public health expenditure is also among the highest, estimated at US$ 360,28 per capita in 2018 (US$ 650 PPP), and its medical workforce includes 163,430 physicians (3.5 per 1,000 inhabitants, the third highest in the country). By contrast, Maranhão state is home to approximately 7.2M people, its per capita income is one-third of São Paulo’s, and only 1% of the population is covered by private health schemes. In 2022, there were 8,743 physicians in Maranhão, that is, 1.22 per 1,000 – the second lowest rate among Brazilian states ^39^.

Our survey was conducted between 16 February and 15 June 2021. The sample was drawn from the nominal listing of physicians registered with Brazil’s Federal Council of Medicine in the two states. The study’s overall sample was composed of 1,183 physicians, consisting of 632 from São Paulo and 551 from Maranhão. The sample was calculated based on the active physicians registered with the Federal Council of Medicine in the two states in 2021 (N=152,511 – 144,852 in São Paulo and 7,659 in Maranhão) and their key demographic characteristics. Proportional stratified sampling was used to replicate the physician distribution by gender, age, state, and residence. A larger proportion of Maranhão’s universe of physicians was selected to allow for a sufficient N for the strata and doctor characteristics of interest. As the two states are very different—in São Paulo there are 30% of all the physicians in Brazil, and in Maranhão a little more than 1%—a proportional sample would have been too small to allow stratifications in Maranhão (see Table 1 in Supplementary Information 2).

**Table 1:**
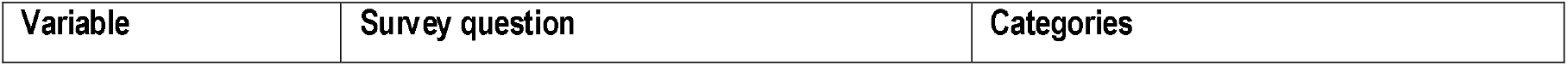
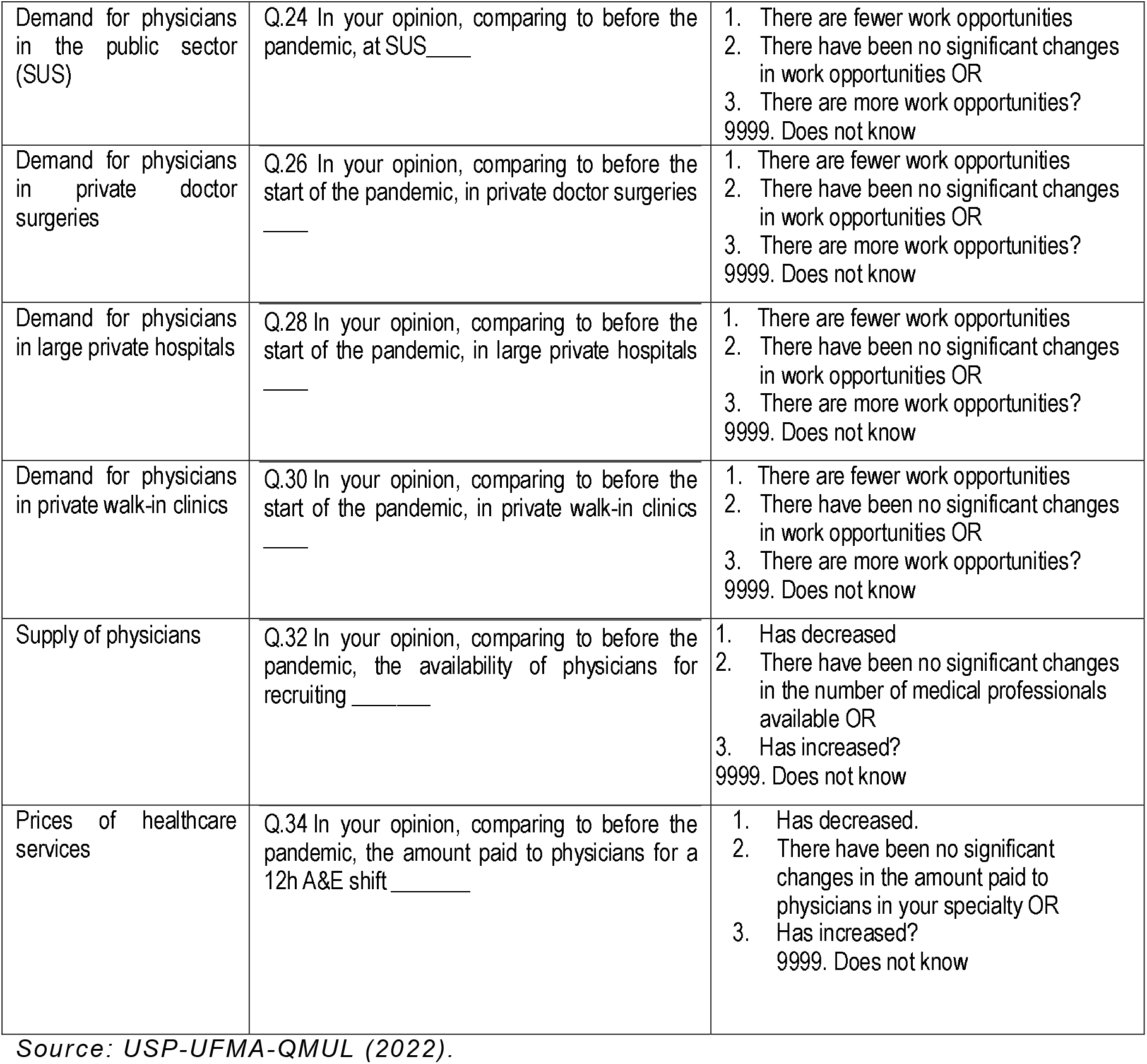
Variables, survey questions, and categories used in the analysis.

| Variable | Survey question | Categories |
| --- | --- | --- |
| Demand for physicians in the public sector (SUS) | Q.24 In your opinion, comparing to before the pandemic, at SUS_____ | 1. There are fewer work opportunities<br>2. There have been no significant changes in work opportunities OR<br>3. There are more work opportunities?<br>9999. Does not know |
| Demand for physicians in private doctor surgeries | Q.26 In your opinion, comparing to before the start of the pandemic, in private doctor surgeries _____ | 1. There are fewer work opportunities<br>2. There have been no significant changes in work opportunities OR<br>3. There are more work opportunities?<br>9999. Does not know |
| Demand for physicians in large private hospitals | Q.28 In your opinion, comparing to before the start of the pandemic, in large private hospitals _____ | 1. There are fewer work opportunities<br>2. There have been no significant changes in work opportunities OR<br>3. There are more work opportunities?<br>9999. Does not know |
| Demand for physicians in private walk-in clinics | Q.30 In your opinion, comparing to before the start of the pandemic, in private walk-in clinics _____ | 1. There are fewer work opportunities<br>2. There have been no significant changes in work opportunities OR<br>3. There are more work opportunities?<br>9999. Does not know |
| Supply of physicians | Q.32 In your opinion, comparing to before the pandemic, the availability of physicians for recruiting _____ | 1. Has decreased<br>2. There have been no significant changes in the number of medical professionals available OR<br>3. Has increased?<br>9999. Does not know |
| Prices of healthcare services | Q.34 In your opinion, comparing to before the pandemic, the amount paid to physicians for a 12h A&E shift _____ | 1. Has decreased.<br>2. There have been no significant changes in the amount paid to physicians in your specialty OR<br>3. Has increased?<br>9999. Does not know |

Source: USP-UFMA-QMUL (2022).

The survey was carried out by a specialised research institute (Datafolha Research Institute), under the technical supervision of the academic researchers. Primary data was collected via a telephone survey conducted in Portuguese by Datafolha data collectors, which included a field coordinator, experienced interviewers, and administrative staff responsible for checking missing data. Sample size calculations, sample selection, questionnaire design, substitution control, database assembly, and data analysis were performed by the authors of this paper.

The original survey received approval from the Research Ethics Committees of the Federal University of Maranhão, Brazil (CEP UFMA 3.051.875) and from the Faculty of Medicine of the University of São Paulo, Brazil (CEP FMUSP 3.136.269).

#### Variables and data analysis

The interviews consisted of a 30-minute telephone questionnaire, containing 30 questions ranging from multiple, closed questions to interdependently concatenated and semi-open questions. The specific variables for this secondary analysis were constructed from the questions below from the Survey Questionnaire’s Section III – Changes in the Labour Market (see Survey Questionnaire in Annex 1).

For our analysis in this study, the prevalence and 95% CIs of variables related to physicians’ perceptions on changes in job opportunities and availability of doctors were estimated for the two states in their facilities of practice (public hospitals-SUS, private doctor’s surgeries, large private hospitals, and walk-in clinics), and stratified for public-only physicians, private-only ones, and dual practitioners (Table 1).

Statistically significant differences at the 5% confidence level were considered in the absence of overlapping 95% CIs. Prevalence and 95 CIs for such perceptions were also analysed and plotted by physicians’ age groups (24–34; 35–44; 45–59; 60+) and by their specific involvement with COVID-19 services. The database developed in Excel by the Datafolha data collectors was exported to R-Studio version 4.1.3 for statistical treatment ^40^.

## Results

Most doctors in our sample said job opportunities and vacancies in the public sector (SUS) increased during COVID-19 (59.0%, 95% CI 56.1-61.9). This was particularly evident among Maranhão doctors (66.4%, 95% CI 62.3-70.3). Opportunities and vacancies were also said to have increased in large private hospitals with in-patient care capacity (51.4%, 95% CI 48.4-54.4), although not as much as in SUS. Again, such positive perceptions were found to be more pronounced among Maranhão doctors (see Table 2).

**Table 2:**
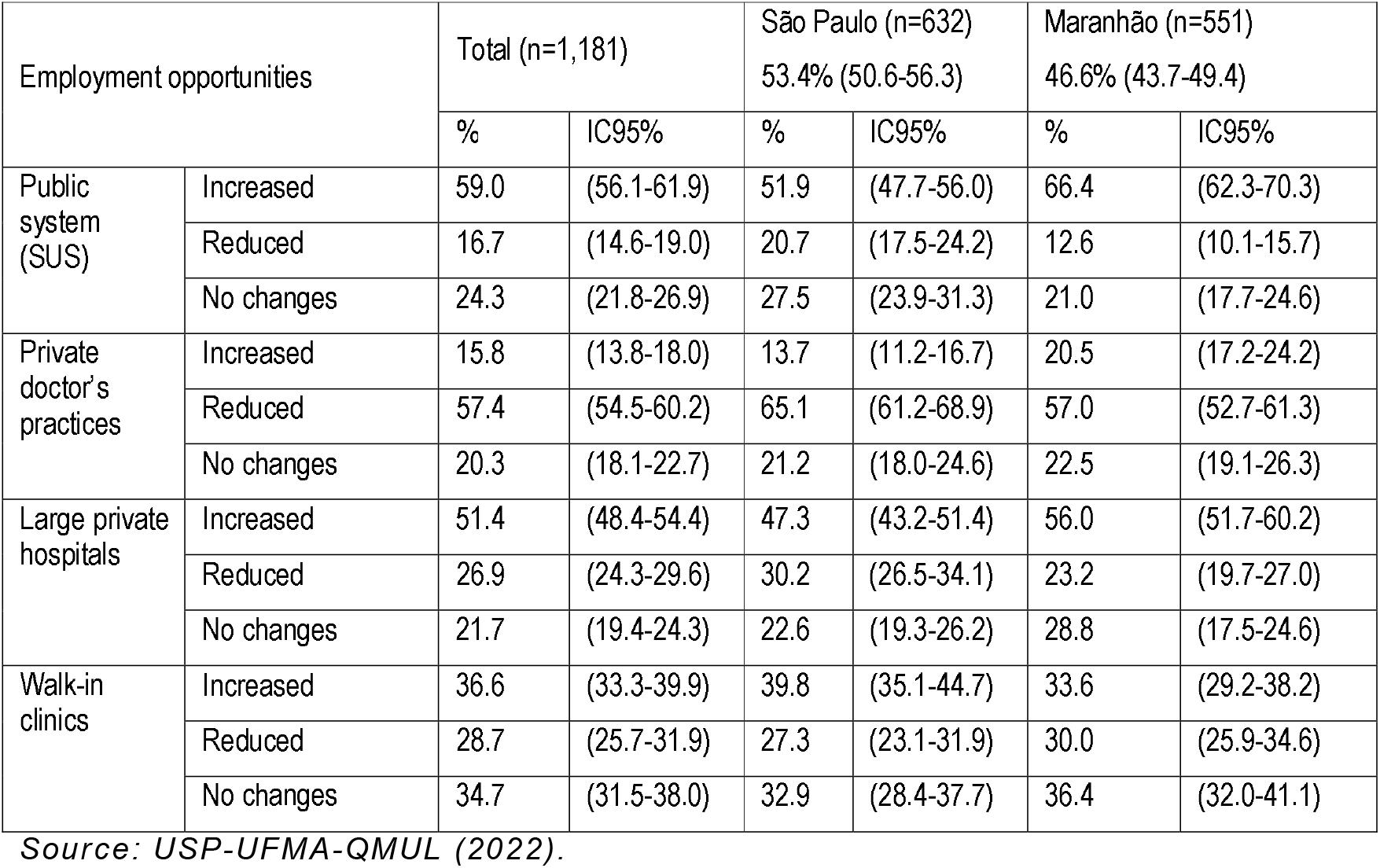
Physicians’ perceptions on employment opportunities in public and private facilities, by sector of employment and state.

For other private sector facilities, perceptions of changing employment opportunities were either negative or neutral, particularly for smaller private doctor’s practices, where 57.4% of doctors declared opportunities to have actually decreased (95% CI 54.5-60.2). Such negative perceptions for smaller private clinics were especially acute in Maranhão (65.1%, 95% CI 61.2-68.9) (Table 2).

These perceptions were confirmed when separating the views of public and private sector doctors, as public doctors declared noticing increased opportunities in their own sector of employment by a larger margin (72.2%, 95% CI 66.1-77.7) and private sector doctors confirmed the reduction of opportunities in private doctor’s practices by 62.7% (95% CI 56.7-68.4). For those doctors working concomitantly in public and private facilities – the dual practitioners – job opportunities increased in the public sector and in large private hospitals (56.4% and 46.3%, respectively), but reduced in smaller doctor’s practices (62.5%, 95 CI 58.8-66.1) and stayed unchanged in walk-in clinics (see Table 2 in Supplementary Information 2).

In regard to the availability of doctors to take up vacancies in specific sectors, there was a slight perception of increased availability among public health physicians (44.8%, 95 CI 38.4-51.4), although this was predominantly driven by the positive perceptions of Maranhão doctors (54.1%, 95 CI 45.7-62.3) – among São Paulo doctors this perception was, in fact, neutral (see Table 3 below). Perceptions of increased availability of doctors were also recorded among dual practitioners in Maranhão (45%, 95 CI 39.7-50.4). For private health physicians, perceptions of positive changes were only significant for Maranhão doctors (46.6%, 95 CI 34.3-59.2).

**Table 3:**
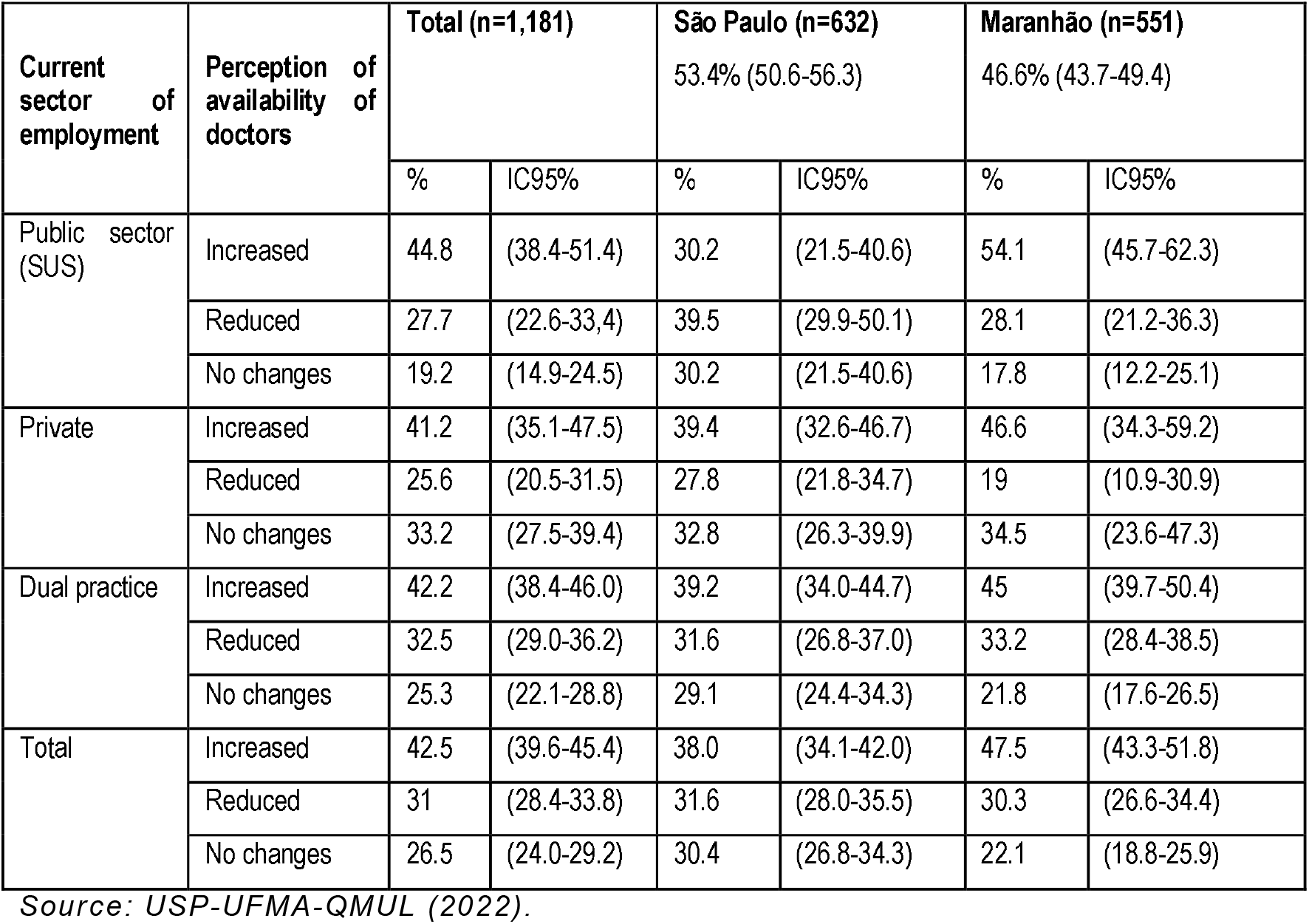
Perceptions on availability of doctors in each sector, by type of current public, private, and dual employment.

| Current sector of employment | Perception of availability of doctors | Total (n=1,181) |  | São Paulo (n=632) |  | Maranhão (n=551) |  |
| --- | --- | --- | --- | --- | --- | --- | --- |
|  |  | % | IC95% | % | IC95% | % | IC95% |
| Public sector (SUS) | Increased | 44.8 | (38.4-51.4) | 30.2 | (21.5-40.6) | 54.1 | (45.7-62.3) |
|  | Reduced | 27.7 | (22.6-33.4) | 39.5 | (29.9-50.1) | 28.1 | (21.2-36.3) |
|  | No changes | 19.2 | (14.9-24.5) | 30.2 | (21.5-40.6) | 17.8 | (12.2-25.1) |
| Private | Increased | 41.2 | (35.1-47.5) | 39.4 | (32.6-46.7) | 46.6 | (34.3-59.2) |
|  | Reduced | 25.6 | (20.5-31.5) | 27.8 | (21.8-34.7) | 19 | (10.9-30.9) |
|  | No changes | 33.2 | (27.5-39.4) | 32.8 | (26.3-39.9) | 34.5 | (23.6-47.3) |
| Dual practice | Increased | 42.2 | (38.4-46.0) | 39.2 | (34.0-44.7) | 45 | (39.7-50.4) |
|  | Reduced | 32.5 | (29.0-36.2) | 31.6 | (26.8-37.0) | 33.2 | (28.4-38.5) |
|  | No changes | 25.3 | (22.1-28.8) | 29.1 | (24.4-34.3) | 21.8 | (17.6-26.5) |
| Total | Increased | 42.5 | (39.6-45.4) | 38.0 | (34.1-42.0) | 47.5 | (43.3-51.8) |
|  | Reduced | 31 | (28.4-33.8) | 31.6 | (28.0-35.5) | 30.3 | (26.6-34.4) |
|  | No changes | 26.5 | (24.0-29.2) | 30.4 | (26.8-34.3) | 22.1 | (18.8-25.9) |

Source: USP-UFMA-QMUL (2022).

Such reported changes in availability of vacancies and doctors, however, were not reflected in the perception of changes in prices; the overwhelming majority of doctors across all sectors (75.1%, 95 CI 72.4-77.6) declared that remuneration for a 12-hour A&E shift stayed broadly unchanged during the pandemic (see Table 3 in Supplementary Information 2).

When disaggregating responses by age groups, younger doctors (aged 24–34) were the ones declaring increased job opportunities, particularly in the public sector (64%, 95 CI 58.1-68.1) (see Figure 1 below).

**Figure 1:**
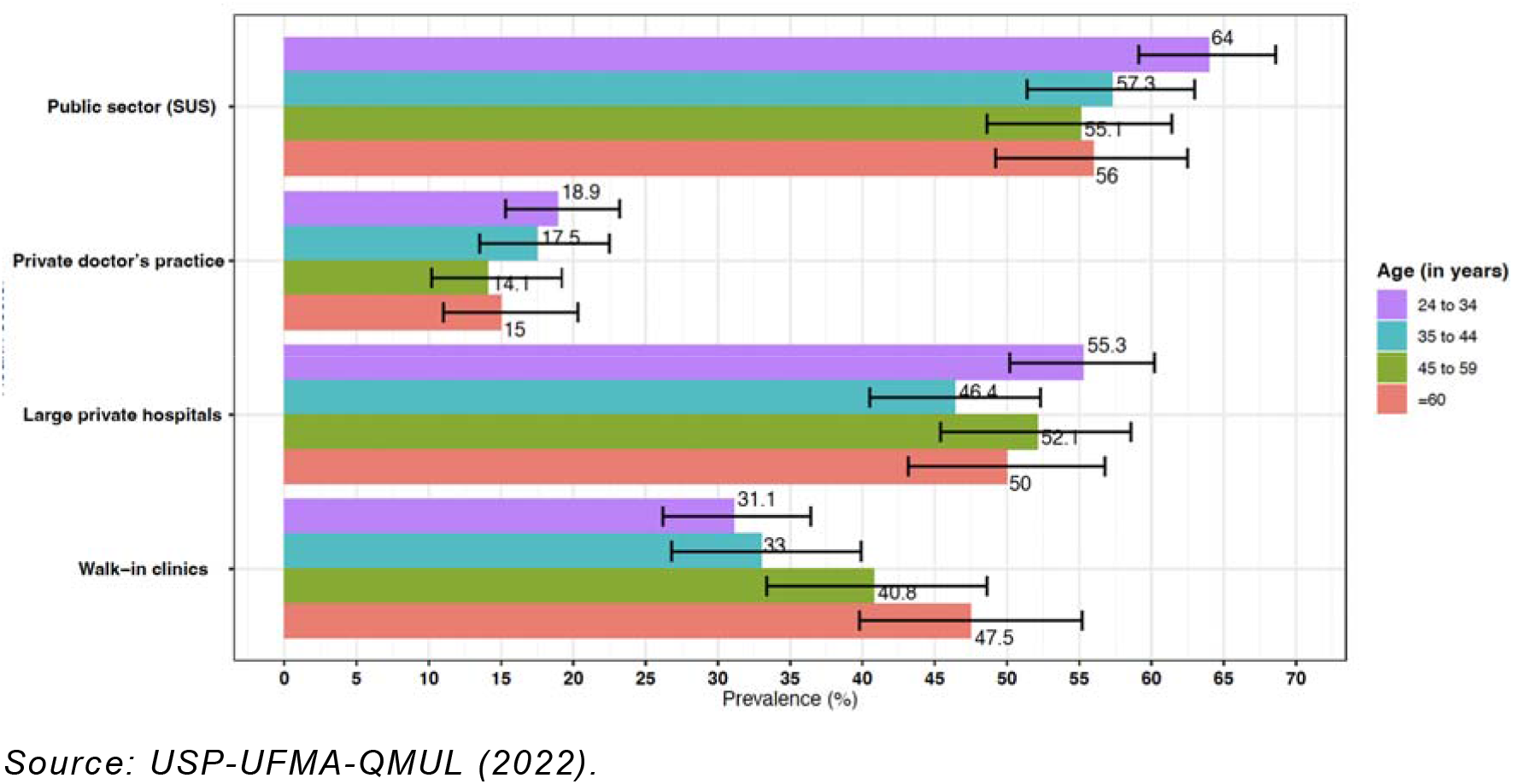
Proportion of doctors reporting increased job opportunities, by age group and type of facility.

In private walk-in clinics, however, it was only the older doctors (>60 years) who reported significantly improved work opportunities (47.5%, 95 CI 39.9-55.1), with all the other age groups reporting either decreased or unchanged employment opportunities (Figure 1).

Doctors working directly with COVID-19 cases generally reported increased opportunities, particularly in public hospitals (65%, 95 CI 62.3-68.4) and in large private ones (55%, 95 CI 51.8-59.1) (see Figure 2). Walk-in clinics were the only exceptions, as in such facilities specialising in working with COVID-19 did not appear to significantly increase job opportunities (35.9%, 95 CI 32.1-40.0).

**Figure 2:**
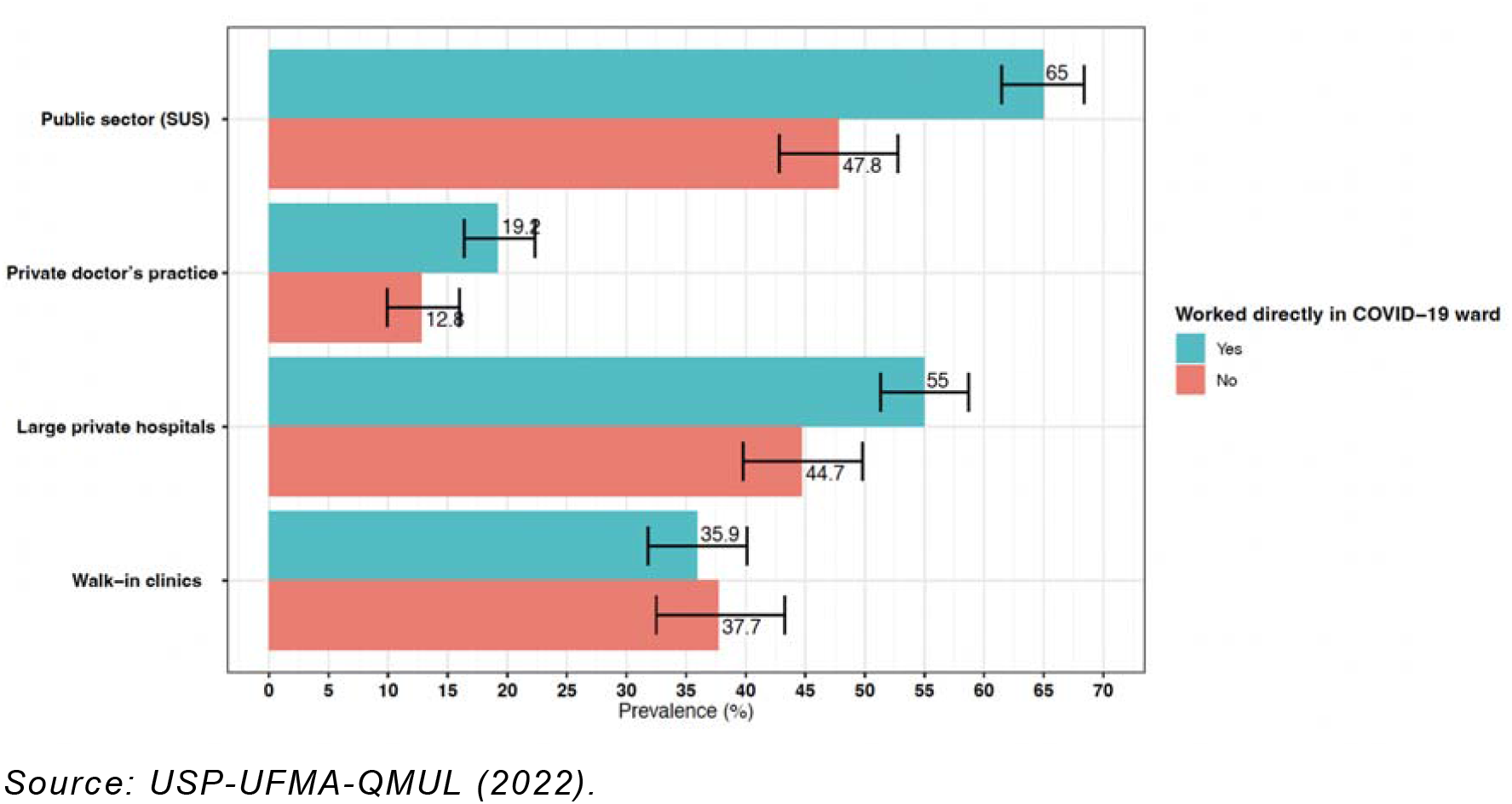
Proportion of doctors declaring increased opportunities, by specialisation in COVID-19 cases and type of facility.

## Discussion

In our survey of labour market perceptions during COVID-19 among physicians in Brazil, we found that most doctors recounted increased job opportunities in the public sector, particularly in Maranhão state. For the private sector, perceptions were mixed, as increased opportunities were reported in large private hospitals but not in smaller practices or walk-in clinics. In regard to the availability of doctors, our survey recorded perceptions of small increases in Maranhão, particularly in the public sector. Remuneration of A&E shifts stayed broadly unchanged. Younger doctors were the ones declaring more job opportunities in public facilities. Older ones reported opportunities in walk-in clinics, particularly in Maranhão. Those doctors working directly with COVID-19 patients saw increases in SUS and large private hospitals, but not elsewhere.

We acknowledge that different doctors across the world experienced COVID-19 in a different way – from the frontline intensive care and infective diseases specialists who found themselves in the eye of the storm, to primary care specialists who transitioned to remote working and telemedicine, to surgeons who simply saw their non-essential procedures cancelled. However, our survey of physicians’ perceptions in Brazil during COVID-19 suggests that job opportunities actually increased in the public sector and in large private hospitals. This is contrast to what was observed for US hospitals during the pandemic ^20^.

Our interpretation is that, in countries like Brazil with publicly funded health systems, resources (and jobs) were proactively redirected toward COVID-19 cases. This would explain why SUS and large private hospitals with in-patient care capacity in Brazil appear to have experienced additional vacancies to meet the increased demand for COVID-19 care. Conversely, smaller private health facilities with mostly outpatient capacity may have temporally suspended some of their operations during the pandemic, in connection with the slow-down of demand for elective procedures. This would be consistent with what was observed in public health systems in European countries ^22^.

Demand and supply for doctors in São Paulo and Maranhão appear to have experienced opposite pandemic effects, with increasing opportunities in the latter state but not in the former. This may be due to the comparatively greater weight of public health services in Maranhão. In São Paulo, the private healthcare sector is very developed ^41^, with an estimated 86% of doctors engaging with it either exclusively or as dual practitioners ^42^. We conclude that the São Paulo health labour market experienced during COVID-19 effects similar to the market in the US, while Maranhão’s displayed features more like the European, Canadian, or Australian markets. Such effects were probably exacerbated by the scarcity of doctors in Maranhão ^43^, who inevitably ended up taking more responsibilities (and risks) in the COVID-19 fight ^36^.

Younger doctors reported increasing job opportunities across the board, particularly in the public sector and in Maranhão. We believe this reflects the decisions taken in Brazil – like in other countries – to deploy younger (and therefore less at risk) cadres to staff COVID-19 services and shelter more senior ones ^36^. This would also be consistent with our findings on the increased opportunities in COVID-19 wards for younger doctors. We interpret the reported increase of opportunities for older doctors in walk-in clinics as an indication that more lucrative parts of the private market would still be primarily accessible to more senior, established physicians, with fewer opportunities for younger doctors ^15^.

Reports of no changes in remuneration for A&E shifts appear to be at odds with standard economic theory that would predict an increase of prices in the presence of increased demand and stable supply of doctors ^44^. On the one hand, we acknowledge that remuneration for 12-hour A&E shifts may not be a suitable bell weather for changes in equilibrium prices for medical services (see the Limitations section below). On the other hand, it may be possible that, in the short run, labour prices for medical services may prove inelastic ^45^, particularly during a pandemic emergency.

Our findings have broader relevance for other countries and future epidemics. We showed that health labour markets do not necessarily shrink during outbreaks, and the impacts will depend on the balance of public and private services within national health systems. Public health systems (and physicians) around the world were a key pillar of policy response to the pandemic, opening new services, performing additional functions, and driving the clinical fight ^46^. This inevitably poses questions on what the role of markets and the private sector should be during a public health emergency ^47^, calling for a re-configuration of the complementarity of public and private functions, with a view to boosting pandemic preparedness in LMICs ^48^.

## Limitations

Our findings need to be interpreted in light of a number of limitations. First, we used physicians’ perceptions of changes in vacancies and prices to gain an insight into the demand and supply of doctors during the pandemic in Brazil. Although workers’ perceptions have been used before in the literature to explore labour market dynamics ^38^, we acknowledge that an examination of employment data would be needed to validate our findings. We also asked our sampled physicians to report on changes that happened in the past two years, which could have been affected by recall bias ^49^.

Secondly, our proxies for demand, supply, and price levels may have left too much room for interpretation, as some of our doctors struggled to distinguish between ‘availability of job opportunities’ and ‘availability of doctors to take up jobs’. Our question on changes in remuneration for A&E shifts was driven by the need to identify a price indicator for medical services that could be known to all the doctors surveyed ^50^. However, we realise that not all doctors in Brazil carry out A&E shifts or necessarily have knowledge of changes in this price.

Finally, we recognise that Maranhão and São Paulo states present very particular configurations of labour market characteristics, organisation of health services, policies, and health workforces ^29,34,39^. Therefore, our findings may not be entirely generalisable to other LMICs.

## Conclusion

Limited evidence exists on health labour markets’ impacts and adaptations during COVID-19, with some literature suggesting a reduction of services. We conducted a secondary analysis of survey data on physicians’ perceptions around changing employment opportunities in one rich and one less developed state in Brazil in 2021, with the objective of gaining insights into health labour markets during epidemics in LMICs.

Most of our sampled doctors noticed increased job opportunities in the public sector, particularly in Maranhão state. For the private sector, perceptions were mixed, as increased opportunities were reported in large private hospitals but not in smaller clinics. Younger doctors perceived an increase of vacancies in public and in large private hospitals, while older ones reported opportunities in walk-in clinics, particularly in Maranhão. Those doctors working directly with COVID-19 patients saw increases in public and large private hospitals, but not elsewhere.

Our findings suggest that health labour markets may not necessarily shrink during epidemics, and that the impacts will depend on the balance of public and private services in national health systems. The complementary roles of health markets and of publicly and privately funded systems during a health emergency should be re-examined, with the objective of improving pandemic preparedness, particularly for LMICs.

## Supporting information

Supplemental material 1 - Survey questionnaire

Supplemental material 2 - Statistical annex

## Data Availability

All data produced in the present study are available upon reasonable request to the authors

## Statements and Declarations

Competing Interests: The authors declare no competing interests.

## Acknowledgments

This study on whose findings this paper is based, received support from the Confap-MRC call for Health Systems Research Networks. Specifically, Bruno de Oliveira was funded by the Fundação de Pesquisa do Estado Maranhão (FAPEMA, Processo COOPI-00709/18) and Coordenação de Aperfeiçoamento de Pessoal de Nível Superior-Brasil (CAPES)—Finance Code 001, to the Graduate Program in Public Health. Giuliano Russo received an award from the NEWTON FUND, MEDICAL RESEARCH COUNCIL (UK), grant reference MR/R022747/1. Mário Scheffer and Alex Cassenote received an award from the Fundação de Amparo à Pesquisa do Estado de São Paulo (FAPESP-Brazil), 2017/50356-7.

## References

1 Wang H, Paulson KR, Pease SA, et al. Estimating excess mortality due to the COVID-19 pandemic: a systematic analysis of COVID-19-related mortality, 2020– 21. The Lancet 2022; 399: 1513–36.

2 Shang Y, Li H, Zhang R. Effects of Pandemic Outbreak on Economies: Evidence From Business History Context. Frontiers in Public Health 2021; 9. https://www.frontiersin.org/articles/10.3389/fpubh.2021.632043 (accessed March 24, 2023).

3 Lee S, Schmidt-Klau D, Verick S. The Labour Market Impacts of the COVID-19: A Global Perspective. Ind J Labour Econ 2020; 63: 11–5.

4 OECD. Labour market developments: The unfolding COVIDl7l19 crisis. Paris: OECD, 2021 DOI:10.1787/7e1e1ad3-en.

5 Russo G, Silva TJ, Gassasse Z, Filippon J, Rotulo A, Kondilis E. The impact of economic recessions on health workers: a systematic review and best-fit framework synthesis of the evidence from the last 50 years. Health Policy Plan 2021; 36: 542–51.

6 Ellis RP, Martins B, Zhu W. Health care demand elasticities by type of service. J Health Econ 2017; 55: 232–43.

7 Staiger DO, Auerbach DI, Buerhaus PI. Registered Nurse Labor Supply and the Recession — Are We in a Bubble? New England Journal of Medicine 2012; 366: 1463–5.

8 Chen A, Sasso AL, Richards MR. Graduating into a downturn: Are physicians recession proof? Health Econ 2018; 27: 223–35.

9 Lavergne MR, Hedden L, Law MR, McGrail K, Ahuja M, Barer M. The impact of the 2008/2009 financial crisis on specialist physician activity in Canada. Health Econ 2018; 27: 1859–67.

10 Galbany-Estragués P, Nelson S. Migration of Spanish nurses 2009-2014. Underemployment and surplus production of Spanish nurses and mobility among Spanish registered nurses: A case study. Int J Nurs Stud 2016; 63: 112–23.

11 Bambra C, Garthwaite K, Copeland A, Barr B. All in it together? Health inequalities, austerity, and the ‘Great Recession’. Oxford University Press, 2015 https://www.oxfordscholarship.com/view/10.1093/acprof:oso/9780198703358.001.0001/acprof-9780198703358-chapter-12 (accessed April 29, 2020).

12 Russo G, Pires CA, Perelman J, Gonçalves L, Barros PP. Exploring public sector physicians’ resilience, reactions and coping strategies in times of economic crisis; findings from a survey in Portugal’s capital city area. BMC Health Serv Res 2017; 17: 207.

13 McPake B, Maeda A, Araújo EC, Lemiere C, El Maghraby A, Cometto G. Why do health labour market forces matter? Bull World Health Organ 2013; 91: 841–6.

14 Scheffler RM, Herbst CH, Lemiere C, Campbell J. Health Labor Market Analyses in Low- and Middle-Income Countries: An Evidence-Based Approach. The World Bank, 2016 DOI:10.1596/978-1-4648-0931-6.

15 McPake B, Scott A, Edoka I. Analyzing Markets for Health Workers: Insights from Labor and Health Economics. The World Bank, 2014 DOI:10.1596/978-1-4648-0224-9.

16 WHO. Health labour market analysis guidebook. Geneva, Switzerland: Health Workforce Department, The World Health Organization, 2021.

17 Causa O, Abendschein M, Luu N, Soldani E, Soriolo C. The post-COVID-19 rise in labour shortages. Paris: OECD, 2022 DOI:10.1787/e60c2d1c-en.

18 Bhandari N, Batra K, Upadhyay S, Cochran C. Impact of COVID-19 on Healthcare Labor Market in the United States: Lower Paid Workers Experienced Higher Vulnerability and Slower Recovery. Int J Environ Res Public Health 2021; 18: 3894.

19 Matta S, Nicholas LH. Changes in Unemployment Among Health Care Workers Following the COVID-19 Pandemic. JAMA 2022; 328: 1639–41.

20 Teasdale B, Schulman KA. Are U.S. Hospitals Still “Recession-proof”? N Engl J Med 2020; 383: e82.

21 Cantor J, Whaley C, Simon K, Nguyen T. US Health Care Workforce Changes During the First and Second Years of the COVID-19 Pandemic. JAMA Health Forum 2022; 3: e215217.

22 Arthur R. Studying the UK job market during the COVID-19 crisis with online job ads. PLOS ONE 2021; 16: e0251431.

23 Šantrić Milićević M, Mandić-Rajčević S, Stevanovic A. Health workers labor market before and during the Covid-19 pandemic: Health sector capacity of Serbia. European Journal of Public Health 2022; 32: ckac131.283.

24 AL-Abrrow H, Al-Maatoq M, Alharbi RK, et al. Understanding employees’ responses to the COVID-19 pandemic: The attractiveness of healthcare jobs. Global Business and Organizational Excellence 2021; 40: 19–33.

25 Shuchman M. Low- and middle-income countries face up to COVID-19. Nature Medicine 2020; 26: 986–8.

26 The World Bank. Brazil Poverty and Equity AssessmentL: Looking Ahead of Two Crises. Washington (DC): The World Bank, 2022 https://documents.worldbank.org/en/publication/documents-reports/documentdetail/099230007062256153/P1746910e33a8407d0b0850b8f0f5bcf18c (accessed March 23, 2023).

27 Flamini V, Toscani F. The Short-Term Impact of COVID-19 on Labor Markets, Poverty and Inequality in Brazil. IMF Working Papers 2021.DOI:https://doi.org/10.5089/9781513571645.001.

28 Nogueira RP. [The socioeconomic crisis and the demand for health professionals in Brazil, 1976-1984]. Bol Oficina Sanit Panam 1988; 104: 572–82.

29 de Oliveira BLCA, Andrietta LS, Reis RS, et al. The Impact of the COVID-19 Pandemic on Physicians’ Working Hours and Earnings in São Paulo and Maranhão States, Brazil. International Journal of Environmental Research and Public Health 2022; 19: 10085.

30 OECD. OECD Reviews of Health Systems: Brazil 2021. OECD, 2021 DOI:10.1787/146d0dea-en.

31 Godoy CV, Silva JB. O Fenômeno de expansão das “clínicas médicas populares” no bairro Centro de Fortaleza/Ceará/Brasil. In: XIV Colóquio Ibérico de Geografia. Fotaleza, Ceará, 2014. http://www.lasics.uminho.pt/conferences/index.php/CEGOT/XIV_ColoquioIbericoGeografia/paper/view/1963 (accessed May 8, 2020).

32 Lapa MR. As clínicas populares como uma alternativa à saúde no Brasil: um estudo de caso em uma clínica popular. 2014. http://www.repositorio.ufc.br/handle/riufc/33277 (accessed May 8, 2020).

33 Gragnolati M, Lindelow M, Couttolenc B. Twenty Years of Health System Reform in Brazil. An Assessment of the Sistema Único de Saúde. Washington, DC: The World Bank, 2013.

34 Andrietta LS, Levi ML, Scheffer MC, Alves MTSS de B e, Oliveira BLCA de, Russo G. The differential impact of economic recessions on health systems in middle-income settings: a comparative case study of unequal states in Brazil. BMJ Global Health 2020; 5: e002122.

35 MRC. How is the current crisis reshaping Brazil’s health system? Strengthening health workforce and provision of services in São Paulo and Maranhão. UK Research and Innovation. 2018. https://gtr.ukri.org/projects?ref=MR%2FR022747%2F1 (accessed June 26, 2019).

36 Russo G, Cassenote A, De Oliveira BLCA, Scheffer M. Demographic and professional risk factors of SARS-CoV-2 infections among physicians in low- and middle-income settings: Findings from a representative survey in two Brazilian states. PLOS Glob Public Health 2022; 2: e0000656.

37 Scheffer M, Cassenote A, de Britto E Alves MTSS, Russo G. The multiple uses of telemedicine during the pandemic: the evidence from a cross-sectional survey of medical doctors in Brazil. Global Health 2022; 18: 81.

38 Dixon JC, Fullerton AS, Robertson DL. Cross-National Differences in Workers’ Perceived Job, Labour Market, and Employment Insecurity in Europe: Empirical Tests and Theoretical Extensions†. European Sociological Review 2013; 29: 1053–67.

39 Russo G, Levi ML, Seabra Soares de Britto E Alves MT, et al. How the ‘plates’ of a health system can shift, change and adjust during economic recessions: A qualitative interview study of public and private health providers in Brazil’s São Paulo and Maranhão states. PLoS One 2020; 15: e0241017.

40 R Core Team — European Environment Agency. R: A language and environment for statistical computing. 2022. https://www.eea.europa.eu/data-and-maps/indicators/oxygen-consuming-substances-in-rivers/r-development-core-team-2006 (accessed March 9, 2022).

41 Cruz JAW, da Cunha MAVC, de Moraes TP, et al. Brazilian private health system: history, scenarios, and trends. BMC Health Services Research 2022; 22: 49.

42 Miotto BA, Guilloux AGA, Cassenote AJF, Mainardi GM, Russo G, Scheffer MC. Physician’s sociodemographic profile and distribution across public and private health care: an insight into physicians’ dual practice in Brazil. BMC Health Serv Res 2018; 18: 299.

43 Scheffer MC, Almeida C, Guilloux AGA, Guerra A, Miotto BA. Demografia médica no Brasil 2023. São Paulo (BRA): Faculdade de Medicina USP, Conselho Federal de Medicina, 2023.

44 Cockx B, Brasseur C. The demand for physician services. Evidence from a natural experiment. J Health Econ 2003; 22: 881–913.

45 BaŞtav L. AN EMPIRICAL STUDY ON THE WAGE AND PRICE STICKINESS OF US ECONOMY (1990-2009) -. Ekonomik Yaklasim 2011; 22: 45–64.

46 Haldane V, De Foo C, Abdalla SM, et al. Health systems resilience in managing the COVID-19 pandemic: lessons from 28 countries. Nat Med 2021; 27: 964–80.

47 Williams OD. COVID-19 and Private Health: Market and Governance Failure. Development 2020; 63: 181–90.

48 David Williams O, Yung KC, Grépin KA. The failure of private health services: COVID-19 induced crises in low- and middle-income country (LMIC) health systems. Global Public Health 2021; 16: 1320–33.

49 Althubaiti A. Information bias in health research: definition, pitfalls, and adjustment methods. J Multidiscip Healthc 2016; 9: 211–7.

50 Lee T, Propper C, Stoye G. Medical Labour Supply and the Production of Healthcare. Fiscal Studies 2019; 40: 621–61.

