## Supplemental material 1 - Survey questionnaire for "What happened to health labour markets during COVID-19? Insights from a survey of medical doctors in Brazil"

**Title:**

**Journal**:

Applied Health Economics and Health Policy

**Authors’ information**

Giuliano Russo (corresponding author), Wolfson Institute of Population Health, Queen Mary University of London, United Kingdom.. ORCID ID: 0000-0002-2716-369X

Bruno Luciano Carneiro Alves de Oliveira, Program in Public Health, Federal University of Maranhão, São Luís 65080-805, Brazil. ORCID ID: 0000-0001-8053-7972

Alex Cassenote, Department of Preventive Medicine, University of São Paulo, Piracicaba 13416-000, Brazil. ORCID ID: 0000-0002-5098-1922

Mário Scheffer. Department of Preventive Medicine, University of São Paulo, Piracicaba 13416-000, Brazil. ORCID ID: 0000-0001-8931-6471

|  | **PM 745123 – SURVEY ON CORONAVIRUS IMPACT ON PHYSICIANS AND LABOUR MARKET IN SÃO PAULO AND MARANHÃO** | | | | | | | | | | | | | | | |  |  |
| --- | --- | --- | --- | --- | --- | --- | --- | --- | --- | --- | --- | --- | --- | --- | --- | --- | --- | --- |
|  |  |  |  |  |  |  |  |  |  |  |  |  |  |  |  |  | **CPD NO.** |  |
|  |  |  |  |  |  |  |  |  |  |  |  |  |  |  |  |  | **CITY NO.** |  |
|  | **INSPECTION** | 1. CHECKED | |  | 2. NO PHONE NUMBER | | | |  | 3. WRONG PHONE NUMBER |  | | 4. RESPONDENT NOT FOUND | | | | **SPOT NO.** |  |
| **CLOSED-ENDED QUESTIONS SCORE** | |  | | | | | | | | | | | | | | QTY: | **SURVEY NO.** |  |
| **OPEN-ENDED QUESTIONS SCORE** | |  | | | | | | | | | | | | | | QTY: | **SURVEY TAKER NO.** |  |
| **TYPE** | | 1. PROBABILISTIC | | | | |  | 2. INTENTIONAL | | | |  | | 3. ENLISTMENT | | | **HOUR START** | **:** |
|  | **INSPECTOR NO.** | |  | | | **CRITIC NO.** | | | |  | **DATE:** | | | | **/ /2020** | | **HOUR END** | **:** |

Good morning/afternoon/evening. My name is _____. I work for Datafolha Research Institute and I am conducting a study, on behalf of ____ **(MENTION FROM LIST: IF SP > SAY USP; IF MA > SAY UFMA)** and the Federal Council of Medicine. Please, could I speak to Dr.______? **(PROCEED WITH THE NAME IN THE LIST. REPEAT FROM START IF THE CALL IS TRANSFERRED.)**

The survey we are conducting is about the medical work in Brazil within the context of the new coronavirus pandemic. The goal of this study is verifying the effects of the COVID-19 pandemic on the medical work in Brazil. This is an anonymous and confidential study that follows the standards of the Research Ethics Committee and the Medical Ethics Committee. Your name and answers will never be identified, and your data will remain stored only for the duration of the study. Do you agree to answer some questions?

1.	Yes	2. No >> **END THE SURVEY**

For quality control purposes, this interview might be recorded.

**GENDER**	1. Male	2. Female

**AGE** What is your age? **(SPONTANEOUS ANSWER, ASK ONLY ONCE)**

|  | **WRITE DOWN** | 1. From 24 to 34 years | 3. From 45 to 59 years |
| --- | --- | --- | --- |
|  |  | 2. From 35 to 44 years | 4. 60 years or older |

**QF.4** Do you currently work in the state of_____ **(SÃO PAULO/ MARANHÃO – MENTION ACCORDING TO THE LIST)**

1. Yes
2. No **>> END THE SURVEY**

**CRM**:

|  | **(GET FROM LIST)** |
| --- | --- |

**QF.3** In what year did you graduate? **(SPONTANEOUS ANSWER, ASK ONLY ONCE)**

|  | **(GET FROM LIST)** |
| --- | --- |

**QF.1** Do you work as a physician or currently works with medicine or health?

1. Yes, full time (exclusivity contract) 🡺 **PROCEED TO Q.36**
2. Yes, half time (partial dedication)
3. Does not work as physician / does not currently work with medicine or health

**QF.2 (IF PF.1= 2 or 3)** Did the decision of (giving partial dedication **IF QF1=2**) or (refrain from dedicating to medical work **IF PF1=3**) occur after March 2020, due to the pandemic?

1. Yes **🡺 PROCEED**
2. No **🡺 THANK THE RESPONDENT AND END THE SURVEY**

**SET 1 – RESPONDENT CHARACTERIZATION**

**P.36** Have you been vaccinated, or do you intend to get vaccinated against COVID-19? **(SPONTANEOUS ANSWER, ASK ONLY ONCE)**

1. He/she is vaccinated
2. He/she has not been vaccinated, but intends to
3. He/she does not intend to get vaccinated

**(SURVEY TAKER, READ THE FOLLOWING)** **THE COVID-19 PANDEMIC START IN BRAZIL IN MARCH 2020. I WILL ASK SOME QUESTIONS ABOUT YOUR WORK BEFORE AND AFTER THE START OF THE PANDEMIC.**

**P.1** Considering your current medical work, with its routine, format, volume of patients, and working hours, when compared to a usual level of work, before March 2020: **(READ PAUSEDLY UNTIL THE QUESTION MARK)**

1. It has not been impacted by the pandemic, performs the same work
2. Has been impacted, but resumed what used to be done before
3. Still suffers from impacts caused by the pandemic
4. The pandemic brought changes that will be permanently incorporated in his/her work
5. Physician work was interrupted due to the pandemic (is not currently working as a physician) **OR**
6. Retired from/definitively abandoned physician work due to the pandemic?

**Q.2** Do you work, or did you work, on a regular basis, with medicine or health ______ ? **(READ EACH ITEM IN THE TABLE)** With any other area or type of activity that was not mentioned? **(IF YES, WRITE DOWN THE AREA OR ACTIVITY)** **(ENCOURAGED ANSWER AND SINGLE ANSWER PER LINE)**

**Q.3 (FOR EACH Q.2=1)** In this medical activity _____ **(READ THE ITEM)**, do you attend or did you attend, on a regular basis, only to patients with health insurance or private patients; attend/attended only SUS patients – (either at a public, philanthropic facility, Social Organization or another type of facility that provides services to SUS) OR attend /attended to both types of patients, work in both public and private sectors? **(ENCOURAGED ANSWER AND SINGLE ANSWER PER LINE)**

| **SHUFFLE ITEMS** | | **Q.2 TYPE OF MEDICAL PRACTICE** | | **Q.3**  **SECTOR OF WORK** | | |
| --- | --- | --- | --- | --- | --- | --- |
|  |  | **YES** | **NO** | **PUBLIC** | **PRIVATE** | **BOTH** |
| **a.** | With appointments at the office, ward or clinic? | **1** | **2** | **1** | **2** | **3** |
| **b.** | In the handling of diagnostic testing equipment? | **1** | **2** | **1** | **2** | **3** |
| **c.** | With surgeries in a hospital that require hospitalization and general/epidural/spinal anesthesia? | **1** | **2** | **1** | **2** | **3** |
| **d.** | With ward surgeries, without hospitalization and local anesthesia? | **1** | **2** | **1** | **2** | **3** |
| **e.** | Management, direction, service administration and institutional roles? | **1** | **2** | **1** | **2** | **3** |
| **f.** | Teaching or research roles? | **1** | **2** | **1** | **2** | **3** |
| **ALWAYS ASK AS LAST QUESTION** | | | | | | |
| **g.** | Do you work in another sector or type of activity that was not mentioned? What___? | **1** | **2** | **1** | **2** | **3** |

**Q.4 (IF Q.3=2 or 3 IN AT LEAST ONE ITEM)** You said that you work/worked in the private sector, attending to patients with health insurance or private patients. I will read a list of some workplaces in the private sector, and I would like to know in each one of them you work at or used to work at on a regular basis.

Do you work or did you work at ______ **(READ EACH ONE OF THE ITEMS)**? Is there any other place of the private sector that I have not mentioned? **(ENCOURAGED ANSWER AND SINGLE ANSWER PER ITEM)**

**Q.5 (FOR EACH Q4=1)** Approximately how many hours do you currently work at ____ **(READ EACH ITEM)** per week? **(ENCOURAGED ANSWER AND SINGLE ANSWER PER ITEM)**

| **SHUFFLE ITEMS** | | **P.4 PRIVATE SECTOR WORK** | | **P.5 HOURS/ WEEK** |
| --- | --- | --- | --- | --- |
|  |  | **YES** | **NO** | **(WRITE DOWN THE HOURS)**  **9996. Zero hours** |
| **a.** | Own private office or shared with one or more colleagues? | **1** | **2** | **______** |
| **b.** | Private clinic or ward, where you provide care and services? | **1** | **2** | **______** |
| **c.** | Affordable private clinics  *(Example: Dr. Consulta, Cia da Consulta, Doutor Hoje for São Paulo; or Super Clínica, Mega Clínica, Super Clínica, Dr. Saúde for Maranhão)* | **1** | **2** | **______** |
| **e.** | Private hospital? | **1** | **2** | **______** |
| **f.** | Private diagnostic laboratory or private clinical analysis facility? | **1** | **2** | **______** |
| **g.** | Pharmaceutical industry? | **1** | **2** | **______** |
| **h.** | Medical sector of a company? | **1** | **2** | **______** |
| **i.** | Private university? | **1** | **2** | **______** |
| **ALWAYS ASK AS LAST QUESTION** | | | | |
| **k.** | Is there any other place of the private sector that I have not mentioned? **(IF YES)** What? __________ **(WRITE DOWN)** | **1** | **2** | **______** |

**Q.6 (IF Q.3=1 OR 3 IN AT LEAST ONE ITEM)** You said that you work/worked in the public sector, attending to SUS patients or in public services or institutions. I will read a list of some workplaces in the public sector, and I would like to know in which one of them do you work at or used to work at on a regular basis.

Do you work or did you work at ______ **(READ EACH ONE OF THE ITEMS)**? Is there any other place of the private sector that I have not mentioned? **(ENCOURAGED ANSWER AND SINGLE ANSWER PER ITEM)**

**Q.7 (FOR EACH Q.6=1)** Approximately how many hours do you currently work at _____ **(READ EACH ITEM)** per week? **(ENCOURAGED ANSWER AND SINGLE ANSWER PER ITEM)**

| **SHUFFLE ITEMS** | | **Q.6 PUBLIC SECTOR WORK** | | **P.7 HOURS/WEEK** |
| --- | --- | --- | --- | --- |
|  |  | **YES** | **NO** | **(WRITE DOWN HOURS)**  **9996. Zero hours** |
| **a.** | Primary health care, such as: *Basic Health Unit, Health Center, Family Health Program, Family Health Strategy* | **1** | **2** | **______** |
| **b.** | Ward services, such as: *Specialty Ward, AMA, mental health services – CAPs; HIV-Aids services; Hemocentre and Hemotherapy; Occupational Health Services* | **1** | **2** | **______** |
| **c.** | Urgent and emergency services, such as: *First aid services, emergency care, UPA (Emergency Unit), rescue, SAMU (Urgent Medical Assistance Service), pre-hospital care* | **1** | **2** | **______** |
| **d.** | Public hospital, such as: *SUS Hospital or an affiliated hospital that attends to SUS patients*  *(Municipal, State or Federal Hospitals, University Hospitals, Charity Hospitals, Philanthropic Hospitals, Hospitals managed by Social Organization, Foundation or Hospital Authority)* | **1** | **2** | **______** |
| **g.** | Public university or public research institutions? | **1** | **2** | **______** |
| **h.** | Administrative or management services of public nature | **1** | **2** | **______** |
| **ALWAYS ASK AS LAST QUESTION** | | | | |
| **k.** | Is there any other place of the public sector that I have not mentioned? **(IF YES)** What? __________ **(WRITE DOWN)** | **1** | **2** | **______** |

**ASK TO ALL RESPONDENTS**

**Q.8** I will read now some different regimes of employment or provision of medical services and I would like to know in which one of them you currently find yourself ______ **(ENCOURAGED ANSWER AND SINGLE ANSWER PER ITEM)**

**Q.9** Before the pandemic, before March 2020, how was your employment regime ______ **(READ EACH ITEM)**?

| **SHUFFLE ITEMS** | | **Q.8 CURRENT**  **EMPLOYMENT**  **METHOD** | | **Q.9 EMPLOYMENT METHOD BEFORE THE PANDEMIC** | |
| --- | --- | --- | --- | --- | --- |
|  |  | **YES** | **NO** | **YES** | **NO** |
| **A.** | Statutory, civil servant? | 1 | 2 | 1 | 2 |
| **B** | CLT employment? | 1 | 2 | 1 | 2 |
| **C.** | Legal entity – PJ? | 1 | 2 | 1 | 2 |
| **D.** | Partner in a legal entity with other professionals? | 1 | 2 | 1 | 2 |
| **E.** | Cooperative? | 1 | 2 | 1 | 2 |
| **F.** | Freelancer with Self-employment income receipt? | 1 | 2 | 1 | 2 |
| **G.** | Contracts or individual agreements of another nature? | 1 | 2 | 1 | 2 |

**Q.10** I will read some income ranges and I would like to know in what range were you in before the pandemic, before March 2020. Considering all your works and employments related to the practice of medicine that you had, approximately how much did you earn per month? **(ENCOURAGED ANSWER AND SINGLE ANSWER)**

1. Up to R$6,000
2. From R$6,001 to R$ 11,000
3. From R$ 11,001 to R$ 16,000
4. From R$ 16,001 to R$ 21,000
5. From R$ 21,001 to R$ 27,000
6. From R$ 27,001 to R$ 32,000
7. From R$ 32,001 to R$ 40,000
8. R$40,001 or more

9997. Refused to answer **(SPONTANEOUS ANSWER)**

**SET II: CHANGES IN WORK OF RESPONDENTS CAUSED BY COVID-19 (both public and private sectors)**

**Q.11**. Since March 2020, have you worked in activities/services directly related to COVID-19 by attending to infected/sick patients or in activities against the pandemic? **(IF YES)** Are you currently working in activities related to COVID-19? **(SPONTANEOUS AND SINGLE ANSWER)**

1. Have not worked and neither is working in any activity directly related to COVID-19.

**IF YES:**

2. Is currently working in activities related to COVID-19.
3. Has worked in activities directly related to COVID-19 but is no longer working at the moment.

**Q.12. (IF Q.11 = 2 or 3)** In what kind of work, activity or place related to COVID-19 did you work/have you worked? **(ENCOURAGED ANSWER AND SINGLE ANSWER PER ITEM)**

| **SHUFFLE ITEMS** | | **Q.12** | |
| --- | --- | --- | --- |
|  |  | **YES** | **NO** |
| **a.** | In hospital care of patients with COVID in ward or ICU | **1** | **2** |
| **b.** | In the care of suspicious or confirmed cases that do not require hospitalization | **1** | **2** |
| **c.** | In teleconsultation or other non-presential guidance or assistance activities related to COVID | **1** | **2** |
| **d.** | In scientific/academic research on COVID | **1** | **2** |
| **g.** | In health surveillance, committees or other governmental actions related to COVIC | **1** | **2** |
| **ALWAYS ASK AS LAST QUESTION** | | | |
| **h.** | Any other activity? What? _____ **(WRITE DOWN)** | **1** | **2** |

**Q.13** Considering a regular/habitual work week before the start of the pandemic, and your work after March 2020, has your number of worked hours per week increased, remained the same or decreased? **(ENCOURAGED ANSWER AND SINGLE ANSWER PER ITEM)**

1. Increased
2. Remained the same
3. Decreased

**Q.14a (IF Q13=1 )** Approximately in how many weekly hours would you say that your work has increased since the start of the pandemic? **(SPONTANEOUS AND SINGLE ANSWER)**

_______ hours **(WRITE DOWN)	9999.** Does not know

**Q.14b (IF Q13=3)** Approximately in how many weekly hours would you say that your work has decreased since the start of the pandemic? **(SPONTANEOUS AND SINGLE ANSWER)**

_______horas **(WRITE DOWN)	9999.** Does not know

**Q.15** Considering your monthly salary before March 2020 – at the start of the pandemic, has your salary been increased, remained the same or decreased? (**ENCOURAGED AND SINGLE ANSWER)**

1. Increased
2. Remained the same
3. Decreased

**Q.16a (IF Q.15 = 1)** How much, in Reais, has your monthly salary been increased? **(ENCOURAGED AND SINGLE ANSWER)**

**(WRITE DOWN)**

**R$**

9997. Refuses to answer

9999. Does not know

**P.16b (SE P.15= 3)** How much, in Reais, has your monthly salary been decreased? **(ENCOURAGED AND SINGLE ANSWER)**

**(WRITE DOWN)**

**R$**

9997. Refuses to answer

9999. Does not know

**P.18** During the pandemic, how much do you think would be a fair amount for one hour of overtime work, in Reais? **(SPONTANEOUS AND SINGLE ANSWER)**

**(WRITE DOWN)**

**R$**

9997. Refuses to answer

9999. Does not know

**Q.19** Were you diagnosed with COVID-19 at any moment? **(IF YES)** Were you asymptomatic, with mild symptoms or did you require hospitalization and/or ICU? **(ENCOURAGED AND SINGLE ANSWER)**

1. Yes, I was asymptomatic
2. Yes, I had mild symptoms
3. Yes, I required hospitalization and/or ICU.
4. I have not been diagnosed with COVID-19.

**Q.20 (IF Q.19 ≠ 4)** How many working days did you miss due to the COVID-19 diagnosis, up until now? **(SPONTANEOUS AND SINGLE ANSWER)**

**(WRITE DOWN)**

9996. None

9999. Does not know

**Q.21.** Do you_______ **(READ EACH ITEM)? (ENCOURAGED AND SINGLE ANSWER)**

| **SHUFFLE ITEMS** | | **Q.22** | |
| --- | --- | --- | --- |
|  |  | **YES** | **NO** |
| **a.** | Carry out appointments and guidance to patients by telemedicine? | **1** | **1** |
| **b.** | Have work meetings by telemedicine? | **1** | **2** |
| **c.** | Have case discussions with colleagues by telemedicine? | **1** | **2** |
| **d.** | Perform prescriptions, certificates or reports by remote methods or telemedicine | **1** | **2** |
| **g.** | Do you prepare/annotate electronic medical records by telemedicine? | **1** | **2** |
| **h.** | Receive medical qualification or training by telemedicine | **1** | **2** |
| **ALWAYS ASK AS LAST QUESTION** | | | |
| **i.** | Perform any other activity by telemedicine? What? _____ **(WRITE DOWN)** | **1** | **2** |

**Q.22 (IF Q.21=1 IN AT LEAST OE ITEM)** Considering your relationship with telemedicine/remote consultation, you: **(READ UNTIL THE QUESTION MARK)** **(ENCOURAGED AND SINGLE ANSWER)**

1. Had already been using this resource before the pandemic and kept using them,
2. Had never used this resource, but started using it due to the pandemic **OR**
3. Used the resource before, but no longer uses it since the beginning of the pandemic?
4. Had already used telemedicine occasionally to receive qualification, but has never used it with patients (SPONTANEOUS ANSWER)

**Q.23 (IF Q.22 = 1 or 2)** Currently, in your professional activities, how many hours do you dedicate yourself on a weekly basis to digital platforms/telemedicine/remote consultation? **(SPONTANEOUS AND SINGLE ANSWER)**

**(ATTENTION, SURVEY TAKER, USE THE BOX WITH MINUTES ONLY IF THE RESPONDENT SPONTANEOUSLY SAYS THE TIME IN MINUTES AND IF HE/SHE MENTIONS LESS THAN 60 MINUTES)**

**(WRITE DOWN in hours)**

**(WRITE DOWN in minutes)**

**(SPONTANEOUS ANSWER)**

9999. Does not know

**SET III: CHANGES IN THE MEDICAL LABOR MARKET**

**Q.24** In your opinion, comparing to before the pandemic, at SUS____ **(READ UNTIL THE QUESTION MARK – ENCOURAGED AND SINGLE ANSWER)**

1. There are fewer work opportunities
2. There have been no significant changes in work opportunities **OR**
3. There are more work opportunities**?**

9999. Does not know

**Q.25a (IF Q.24=3)** Using a score from one to ten, how much do you think that the work opportunities at SUS have been increased when compared to before the start of the pandemic?

| **ZERO** |  |  |  |  |  |  |  |  |  | **TEN** | **DOES NOT KNOW** |
| --- | --- | --- | --- | --- | --- | --- | --- | --- | --- | --- | --- |
| 9996 | 1 | 2 | 3 | 4 | 5 | 6 | 7 | 8 | 9 | 10 | 9999 |

**Q.25b (IF Q.24=1)** Using a score from one to ten, how much do you think that the work opportunities at SUS have been decreased when compared to before the start of the pandemic?

| **ZERO** |  |  |  |  |  |  |  |  |  | **TEN** | **DOES NOT KNOW** |
| --- | --- | --- | --- | --- | --- | --- | --- | --- | --- | --- | --- |
| 9996 | 1 | 2 | 3 | 4 | 5 | 6 | 7 | 8 | 9 | 10 | 9999 |

**Q.26** In your opinion, comparing to before the start of the pandemic, at the private offices ____  **(READ UNTIL THE QUESTION MARK – ENCOURAGED AND SINGLE ANSWER)**

1. There are fewer work opportunities
2. There have been no significant changes in work opportunities **OR**
3. There are more work opportunities**?**
9999. Does not know

**Q.27a (IF Q.26=3)** Using a score from one to ten, how much do you think that the work opportunities at private offices have been increased when compared to before the start of the pandemic?

| **ZERO** |  |  |  |  |  |  |  |  |  | **TEM** | **DOES NOT KNOW** |
| --- | --- | --- | --- | --- | --- | --- | --- | --- | --- | --- | --- |
| 9996 | 1 | 2 | 3 | 4 | 5 | 6 | 7 | 8 | 9 | 10 | 9999 |

**Q.27b (IF Q.27=1)** Using a score from one to ten, how much do you think that the work opportunities at private offices have been decreased when compared to before the start of the pandemic?

| **ZERO** |  |  |  |  |  |  |  |  |  | **TEN** | **DOES NOT KNOW** |
| --- | --- | --- | --- | --- | --- | --- | --- | --- | --- | --- | --- |
| 9996 | 1 | 2 | 3 | 4 | 5 | 6 | 7 | 8 | 9 | 10 | 9999 |

**Q.28** In your opinion, comparing to before the start of the pandemic, at the private hospitals ____  **(READ UNTIL THE QUESTION MARK – ENCOURAGED AND SINGLE ANSWER)**

1. There are fewer work opportunities
2. There have been no significant changes in work opportunities **OR**
3. There are more work opportunities**?**

9999. Does not know

**Q.29a (IF Q.28=3)** Using a score from one to ten, how much do you think that the work opportunities at private hospitals have been increased when compared to before the start of the pandemic?

| **ZERO** |  |  |  |  |  |  |  |  |  | **TEN** | **DOES NOT KNOW** |
| --- | --- | --- | --- | --- | --- | --- | --- | --- | --- | --- | --- |
| 9996 | 1 | 2 | 3 | 4 | 5 | 6 | 7 | 8 | 9 | 10 | 9999 |

**Q.29b (IF Q.28=1)** Using a score from one to ten, how much do you think that the work opportunities at private hospitals have been decreased when compared to before the start of the pandemic?

| **ZERO** |  |  |  |  |  |  |  |  |  | **TEN** | **DOES NOT KNOW** |
| --- | --- | --- | --- | --- | --- | --- | --- | --- | --- | --- | --- |
| 9996 | 1 | 2 | 3 | 4 | 5 | 6 | 7 | 8 | 9 | 10 | 9999 |

**Q.30** In your opinion, comparing to before the start of the pandemic, at the affordable private clinics ____ **(READ UNTIL THE QUESTION MARK – ENCOURAGED AND SINGLE ANSWER)**

1. There are fewer work opportunities
2. There have been no significant changes in work opportunities **OR**
3. There are more work opportunities**?**

9999. Does not know

**Q.31a (IF Q.30=3)** Using a score from one to ten, how much do you think that the work opportunities at affordable private clinics have been increased when compared to before the start of the pandemic?

| **ZERO** |  |  |  |  |  |  |  |  |  | **TEN** | **DOES NOT KNOW** |
| --- | --- | --- | --- | --- | --- | --- | --- | --- | --- | --- | --- |
| 9996 | 1 | 2 | 3 | 4 | 5 | 6 | 7 | 8 | 9 | 10 | 9999 |

**Q.31b (IF Q.30=1)** Using a score from one to ten, how much do you think that the work opportunities at affordable private clinics have been decreased when compared to before the start of the pandemic?

| **ZERO** |  |  |  |  |  |  |  |  |  | **TEN** | **DOES NOT KNOW** |
| --- | --- | --- | --- | --- | --- | --- | --- | --- | --- | --- | --- |
| 9996 | 1 | 2 | 3 | 4 | 5 | 6 | 7 | 8 | 9 | 10 | 9999 |

**Q.32** In your opinion, comparing to before the pandemic, the availability of physicians for hiring _______ **(READ UNTIL THE QUESTION MARK – ENCOURAGED AND SINGLE ANSWER)**

1. Has been decreased
2. There have been no significant changes in the number of medical professionals available **OR**
3. Has been increased**?**

9999. Does not know

**Q.33a (IF Q.32=3)** Using a score from one to ten, how much do you think that the availability of medical professionals for hiring has been increased, when compared to before the start of the pandemic?

| **ZERO** |  |  |  |  |  |  |  |  |  | **TEN** | **DOES NOT KNOW** |
| --- | --- | --- | --- | --- | --- | --- | --- | --- | --- | --- | --- |
| 9996 | 1 | 2 | 3 | 4 | 5 | 6 | 7 | 8 | 9 | 10 | 9999 |

**Q.33b (IF Q.32=1)** Using a score from one to ten, how much do you think that the availability of medical professionals for hiring has been decreased, when compared to before the start of the pandemic?

| **ZERO** |  |  |  |  |  |  |  |  |  | **TEN** | **DOES NOT KNOW** |
| --- | --- | --- | --- | --- | --- | --- | --- | --- | --- | --- | --- |
| 9996 | 1 | 2 | 3 | 4 | 5 | 6 | 7 | 8 | 9 | 10 | 9999 |

**Q.34** In your opinion, comparing to before the pandemic, the amount paid to physicians on duty in your specialty _______ **(READ UNTIL THE QUESTION MARK – ENCOURAGED AND SINGLE ANSWER)**

1. Has been decreased
2. There have been no significant changes in the amount paid to physicians in your specialty **OR**
3. Has been increased**?**

9999. Does not know

**Q.35a (IF Q34=3)** In terms of a percentage, how much do you think that the amount paid to physicians on duty in your specialty has been increased, when compared to before the pandemic? **(SINGLE ANSWER)**

**(WRITE DOWN – 0% to 100%)**

9999. Does not know

**Q.35b (IF Q34=1)** In terms of a percentage, how much do you think that the amount paid to physicians on duty in your specialty has been decreased, when compared to before the pandemic? **(SINGLE ANSWER)**

**(WRITE DOWN– 0% to 100%)**

9999. Does not know

**AGRADEÇA E ENCERRE**

I declare hereby that:

1. this survey was conducted in accordance with the instructions of the field supervision;

2. the information in this survey were correctly written down and they accurately correspond to the declarations of the respondent;

3. I am aware that the material collected by me is being or will be checked for quality control purposes;

4. I am aware that I must keep the confidentiality of the collected information;

5. I shall not reproduce this survey nor the information in it for my own use or third-party use.

NAME : ___________________________________________________________________________________

SIGNATURE: __________________________ID. Doc. No.: __________________Date: ____/ ___ / ___
