## Supplemental material 2 - Statistical annex for "What happened to health labour markets during COVID-19? Insights from a survey of medical doctors in Brazil"

**Title:**

**Journal**:

Applied Health Economics and Health Policy

**Authors’ information**

Giuliano Russo (corresponding author), Wolfson Institute of Population Health, Queen Mary University of London, United Kingdom.. ORCID ID: 0000-0002-2716-369X

Bruno Luciano Carneiro Alves de Oliveira, Program in Public Health, Federal University of Maranhão, São Luís 65080-805, Brazil. ORCID ID: 0000-0001-8053-7972

Alex Cassenote, Department of Preventive Medicine, University of São Paulo, Piracicaba 13416-000, Brazil. ORCID ID: 0000-0002-5098-1922

Mário Scheffer. Department of Preventive Medicine, University of São Paulo, Piracicaba 13416-000, Brazil. ORCID ID: 0000-0001-8931-6471

Table 1: Physicians sample, but location and socio-economic characteristics

| Physicians’ sociodemographic characteristics | Proportion of total (n = 1,183) | São Paulo (n = 632) 53.4% | Maranhão (n = 551) 46.6% |
| --- | --- | --- | --- |
| Gender |  |  |  |
| Male | 56.2 | 54.1 | 58.6 |
| Female | 43.8 | 45.9 | 41.4 |
| Age |  |  |  |
| 24 to 34 | 34.1 | 34.3 | 33.9 |
| 35 to 44 | 24.5 | 20.7 | 28.9 |
| 45 to 59 | 20.4 | 22.3 | 18.1 |
| ≥60 | 21.0 | 22.6 | 19.1 |
| Geographical location of deployment |  |  |  |
| Rural areas (Interior) | 50.5 | 54.9 | 45.6 |
| Urban areas around capital cities | 49.5 | 45.1 | 54.5 |
| Health sector of deployment |  |  |  |
| Exclusively public | 19.6 | 14.2 | 25.8 |
| Exclusively private | 22.1 | 31.5 | 11.3 |
| Dual practice | 58.3 | 54.3 | 63.0 |
| Working directly with COVID-19 patients |  |  |  |
| Yes | 63.4 | 59.2 | 68.2 |
| No | 36.6 | 40.8 | 31.8 |

Table 2: Physicians' perceptions on employment opportunities in public and private facilities, by State

| **Employment opportunities** | | **Total (n=1,181)** | | **São Paulo (n=632)**  **53.4% (50.6-56.3)** | | **Maranhão (n=551)**  **46.6% (43.7-49.4)** | |
| --- | --- | --- | --- | --- | --- | --- | --- |
|  |  | % | IC95% | % | IC95% | % | IC95% |
| **Public sector** |  |  |  |  |  |  |  |
| SUS | Increased | 72.2 | (66.1-77.7) | 67.4 | (57.0-76.4) | 75.2 | (67.4-81.6) |
|  | Reduced | 11.9 | (8.3-16.8) | 18.6 | (11.8-28.1) | 7.8 | (4.4-13.4) |
|  | No changes | 15.9 | (11.7-21.2) | 14.0 | (8.2-22.8) | 17.0 | (11.7-24.1) |
| Private doctor’s practice | Increased | 18.8 | (13.9-25.0) | 13.0 | (7.0-23.0) | 22.2 | (15.6-30.6) |
|  | Reduced | 55.4 | (48.2-62.3) | 65.2 | (53.4-75.4) | 49.6 | (40.7-58.5) |
|  | No changes | 25.8 | (20.1-32.5) | 21.7 | (13.6-32.8) | 28.2 | (20.8-37.0) |
| Large private hospitals | Increased | 70.2 | (63.5-76.1) | 68.9 | (57.7-78.3) | 71.0 | (62.4-78.2) |
|  | Reduced | 12.6 | (8.7-18.0) | 14.9 | (8.5-24.7) | 11.3 | (6.9-18.1) |
|  | No changes | 17.2 | (12.6-23.0) | 16.2 | (9.5-26.2) | 17.7 | (12.0-25.4) |
| Walk-in private clinics | Increased | 40.9 | (34.0-48.2) | 45.9 | (34.0-58.3) | 38.3 | (30.1-47.3) |
|  | Reduced | 21.0 | (15.7-27.5) | 21.3 | (12.9-33.1) | 20.8 | (14.5-28.9) |
|  | No changes | 38.1 | (31.4-45.4) | 32.8 | (22.3-45.3) | 40.8 | (32.5-49.8) |
| **Private sector** |  |  |  |  |  |  |  |
| SUS | Increased | 53.1 | (46.4-59.6) | 47.4 | (39.7-55.3) | 67.8 | (55.1-78.3) |
|  | Reduced | 17.8 | (13.3-23.5) | 19.5 | (14.0-26.4) | 13.6 | (7.0-24.5) |
|  | No changes | 29.1 | (23.4-35.5) | 33.1 | (26.2-40.9) | 18.6 | (10.7-30.4) |
| Private doctor’s practice | Increased | 12.2 | (8.7-16.7) | 9.2 | (5.9-14.0) | 22.0 | (13.4-34.1) |
|  | Reduced | 62.7 | (56.7-68.4) | 65.8 | (58.9-72.1) | 52.5 | (40.0-64.7) |
|  | No changes | 25.1 | (20.2-30.8) | 25.0 | (19.5-31.5) | 25.4 | (16.1-37.8) |
| Large private hospitals | Increased | 49.6 | (43.3-55.9) | 45.2 | (38.0-52.6) | 62.7 | (50.0-73.9) |
|  | Reduced | 24.2 | (19.1-30.0) | 26.0 | (20.1-32.9) | 18.6 | (10.7-30.4) |
|  | No changes | 26.3 | (21.1-32.2) | 28.8 | (22.6-35.9) | 18.6 | (10.7-30.4) |
| Walk-in private clinics | Increased | 37.1 | (30.3-44.5) | 36.4 | (28.6-45.0) | 39.1 | (26.4-53.5) |
|  | Reduced | 28.6 | (22.4-35.7) | 29.5 | (22.3-37.8) | 26.1 | (15.6-40.3) |
|  | No changes | 34.3 | (27.7-41.6) | 34.1 | (26.5-42.6) | 34.8 | (22.7-49.2) |
| **Dual practice** |  |  |  |  |  |  |  |
| SUS | Increased | 56.4 | (52.6-60.1) | 49.8 | (44.4-55.3) | 62.5 | (57.3-67.5) |
|  | Reduced | 18.0 | (15.3-21.1) | 21.8 | (17.6-26.6) | 14.5 | (11.1-18.6) |
|  | No changes | 25.6 | (22.4-29.1) | 28.3 | (23.7-33.5) | 23.0 | (18.8-27.8) |
| Private doctor’s practice | Increased | 18.1 | (15.4-21.2) | 16.6 | (12.9-21.0) | 19.6 | (15.7-24.2) |
|  | Reduced | 62.5 | (58.8-66.1) | 64.7 | (59.4-69.7) | 60.4 | (55.1-24.5) |
|  | No changes | 19.3 | (16.5-22.5) | 18.7 | (14.9-23.3) | 19.9 | (16.0-24.5) |
| Large private hospitals | Increased | 46.3 | (42.4-50.1) | 43.4 | (38.0-48.9) | 49.1 | (43.7-54.5) |
|  | Reduced | 32.2 | (28.7-36.0) | 36.1 | (31.0-41.5) | 28.5 | (23.9-33.7) |
|  | No changes | 21.5 | (18.5-24.8) | 20.6 | (16.5-25.4) | 22.4 | (18.2-27.2) |
| Walk-in private clinics | Increased | 34.6 | (30.4-39.1) | 40.1 | (33.6-47.0) | 30.4 | (25.1-36.2) |
|  | Reduced | 31.8 | (27.7-36.2) | 27.7 | (22.0-34.3) | 35.0 | (29.5-41.0) |
|  | No changes | 33.5 | (29.4-38.0) | 32.2 | (26.1-38.9) | 34.6 | (29.1-40.6) |
| **Total** |  |  |  |  |  |  |  |
| SUS | Increased | 59.0 | (56.1-61.9) | 51.9 | (47.7-56.0) | 66.4 | (62.3-70.3) |
|  | Reduced | 16.7 | (14.6-19.0) | 20.7 | (17.5-24.2) | 12.6 | (10.1-15.7) |
|  | No changes | 24.3 | (21.8-26.9) | 27.5 | (23.9-31.3) | 21.0 | (17.7-24.6) |
| Private doctor’s practice | Increased | 15.8 | (13.8-18.0) | 13.7 | (11.2-16.7) | 20.5 | (17.2-24.2) |
|  | Reduced | 57.4 | (54.5-60.2) | 65.1 | (61.2-68.9) | 57.0 | (52.7-61.3) |
|  | No changes | 20.3 | (18.1-22.7) | 21.2 | (18.0-24.6) | 22.5 | (19.1-26.3) |
| Hospitais Large private hospitals | Increased | 51.4 | (48.4-54.4) | 47.3 | (43.2-51.4) | 56.0 | (51.7-60.2) |
|  | Reduced | 26.9 | (24.3-29.6) | 30.2 | (26.5-34.1) | 23.2 | (19.7-27.0) |
|  | No changes | 21.7 | (19.4-24.3) | 22.6 | (19.3-26.2) | 28.8 | (17.5-24.6) |
| Walk-in private clinics | Increased | 36.6 | (33.3-39.9) | 39.8 | (35.1-44.7) | 33.6 | (29.2-38.2) |
|  | Reduced | 28.7 | (25.7-31.9) | 27.3 | (23.1-31.9) | 30.0 | (25.9-34.6) |
|  | No changes | 34.7 | (31.5-38.0) | 32.9 | (28.4-37.7) | 36.4 | (32.0-41.1) |

Source: USP-UFMA-QMUL (2022)

Table 3: Perceptions on changes in remuneration for 12h A&E shift

| Remuneration per shift | | Total (n=1,181) | | São Paulo (n=632)  53.4% (50.6-56.3) | | Maranhão (n=551)  46.6% (43.7-49.4) | |
| --- | --- | --- | --- | --- | --- | --- | --- |
|  |  | % | IC95% | % | IC95% | % | IC95% |
| Público | Increased | 15.7 | (11.5-21.2) | 14.8 | (8.7-24.1) | 16.3 | (11.0-23.4) |
|  | Reduced | 7.9 | (5.0-12.2) | 7.4 | (3.4-15.2) | 8.2 | (4.6-14.0) |
|  | No changes | 76.4 | (70.3-81.6) | 77.8 | (67.7-85.5) | 75.6 | (67.7-82.0) |
| Privado | Increased | 8.6 | (5.5-13.2) | 8.8 | (5.3-14.2) | 8.0 | (3.2-18.8) |
|  | Reduced | 10.5 | (7.0-15.4) | 10.7 | (6.8-16.5) | 10.0 | (4.4-21.4) |
|  | No changes | 80.9 | (75.0-85.6) | 80.5 | (73.7-85.9) | 82.0 | (69.2-90.2) |
| Dupla prática | Increased | 12.4 | (10.0-15.1) | 12.8 | (9.6-16.9) | 11.9 | (8.8-16.0) |
|  | Reduced | 14.9 | (12.3-17.8) | 16.2 | (12.6-20.6) | 13.5 | (10.2-17.7) |
|  | No changes | 72.8 | (69.2-76.1) | 71.0 | (65.8-75.7) | 74.5 | (69.5-79.0) |
| Total | Increased | 12.3 | (10.5-14.4) | 11.9 | (9.5-14.9) | 12.7 | (10.1-15.9) |
|  | Reduced | 12.6 | (10.7-14.7) | 13.4 | (10.8-16.4) | 11.7 | (9.2-14.8) |
|  | No changes | 75.1 | (72.4-77.6) | 74.7 | (70.9-78.1) | 75.5 | (71.6-79.1) |

Source: USP-UFMA-QMUL (2022)
